# Strengthening Childhood Cancer Care in Nepal (DECAN-CHILD): Protocol for Evaluating Quality, Financial Burden and Health System Readiness Through a Prospective Cohort Study

**DOI:** 10.64898/2026.10.23.26366300

**Authors:** Deependra Singh, Ramhari Chapagain, Shree Krishna Shrestha, Krishna Prasad Paudel, Dipendra Raman Singh, Samikshya Adhikari, Sudhir Sapkota, Sani Sipai, Ujjwal Chalise, Ritu Lamichhane, Sucharita Tuladhar, Anjali Pandit, Krishna Sagar Sharma, Niva Tiwari, Saroj Yadav, Isabel Arruda-Caycho, Eric Lucas, Richard Muwonge, Catherine Lam, Md Irteja Islam, Partha Basu, Alexandra Martiniuk, Beverley M. Essue

## Abstract

**Background:** Strengthening health systems in low-and middle-income countries (LMICs) like Nepal is essential for equitable, high-quality diagnosis and treatment and improved survival from childhood cancer. This study aims to quantify care intervals and their determinants; assess the quality of care and 1-, 2-, and 3-year survival; quantify the costs and cost burden of childhood cancer treatment from family and health system perspectives; and to assess the capacity of health systems in Nepal to diagnose and treat children with cancer.

**Methods:** This three-year prospective cohort study will enrol and follow consecutive childhood cancer patients aged 0-19 years registered at six tertiary hospitals in Nepal, accounting for approximately 90% of childhood cancer cases diagnosed nationally. After obtaining informed consent and assent, a semi structured questionnaire will be administered to patient/caregiver at baseline and every three months during the first year and every six months in the second and third years. Patient/caregiver data will be supplemented with patients’ medical records kept in the hospital. A national capacity assessment will assess readiness for childhood cancer care in tertiary-level and secondary-level healthcare facilities involved in referral, diagnosis, treatment or shared care. Survival outcomes will be examined using Cox proportional hazards models and random effects models to account for hospital clustering. Out-of-pocket costs will be analysed using descriptive statistics and a two-part regression model to account for zero-inflated cost distributions, with five-year health system cost projections modelled to estimate corresponding rates of catastrophic health expenditure and medical impoverishment.

**Discussion:** This study will generate comprehensive evidence on childhood cancer care, costs, and health system capacity for care in Nepal. Findings will directly inform policy guidance, resource allocation and benefit package design for childhood cancer care in Nepal, inform comparable efforts in other low- and middle-income countries (LMICs), and will contribute to Nepal’s efforts in the WHO Global Initiative for Childhood Cancer towards improved cancer outcomes.

## INTRODUCTION

Childhood cancers have a high cure rate of around 80% in high-income countries (HICs), but less than 30% of children in low- and middle-income countries (LMICs) survive cancer [1,2]. Given that roughly 90% of the world’s children live in LMICs, addressing childhood cancer in these settings is a global health priority [3]. In Nepal, approximately 1-1.5 per 10,000 children aged 0-19 develop cancer annually, equating to 1,100–1,600 new cases expected annually [4,5] with an estimated survival rate of about 20–30% [6]. Survival disparities are largely due to limited access to timely and quality care, delayed diagnoses, treatment abandonment, supportive care or therapeutic intensity, and high financial burden of treatment on families due to financing and financial protection gaps for cancer care [2,7–9]. Only 29% of LMICs report access to essential cancer medicines, compared to 96% of HICs [10,11], and fewer than 30% of children in LMICs receive a timely diagnosis and effective treatment [12].

To address the challenges, in 2018, the World Health Organization (WHO) with support of St. Jude Children’s Research Hospital launched the Global Initiative for Childhood Cancer, aiming to increase global childhood cancer survival to 60% by 2030 [6]. Nepal, a lower-middle income country in South Asia, was identified as a priority country for implementation and systems strengthening support in this initiative [13,14]. The goal of this study is to enhance health systems readiness for delivering a comprehensive and systems-oriented approach to childhood cancer care in Nepal. The research consists of four specific aims that will support a new landscape for childhood cancer and health systems insights in Nepal, with relevance and applicability for similar LMIC settings. To achieve this goal, there are four study objectives:

1. To identify and quantify access, diagnostic, and treatment intervals in childhood cancer care pathways and their determinants.
2. To assess the quality of cancer diagnosis, care and survival at 1, 2 and 3 years following confirmatory diagnosis.
3. To quantify the costs and cost burden of childhood cancer care from the family and health systems perspectives to characterize financial protection for childhood cancer care in Nepal.
4. To assess health facilities’ capacity to diagnose and treat children with cancer.

## METHODS

### Study design and settings

This is a three-year prospective cohort study with a longitudinal survey design and institutional capacity, and readiness assessment. An overview of the study settings, procedures, and workplan are outlined in Figure 1. Six hospitals have been selected for patient recruitment based on the geographic, ethnic, cultural and socio-economic diversity that reflects the national population distribution and predominant service delivery sites [15]. Five hospitals are in Kathmandu Valley, four of which are public hospitals (Bhaktapur Cancer Hospital, Kanti Children’s Hospital, Shree Birendra Hospital, Patan Academy of Health Sciences) delivering care that is either subsidized or free at the point of care, and the privately owned Nepal Cancer Hospital is the largest private providers of child oncology services, where most care is financed through out-of-pocket payments by the patients/caregivers. The sixth hospital, B.P. Koirala Memorial Cancer Hospital, is the largest public oncology center located 160 kilometers south-west of Kathmandu. The public hospitals are actively engaged in the Global Platform for Access to Childhood Cancer Medicines, with a recent Ministry declaration of free care for children with cancer under age 14 years in these facilities [14]. These hospitals provide emergency, outpatient, and inpatient care including chemotherapy, radiotherapy, and surgery. In addition, we will conduct health facility assessment in selected tertiary and secondary hospitals, with geographical and population representation, to examine health facility readiness for providing childhood cancer care services in Nepal.

**Figure 1.**
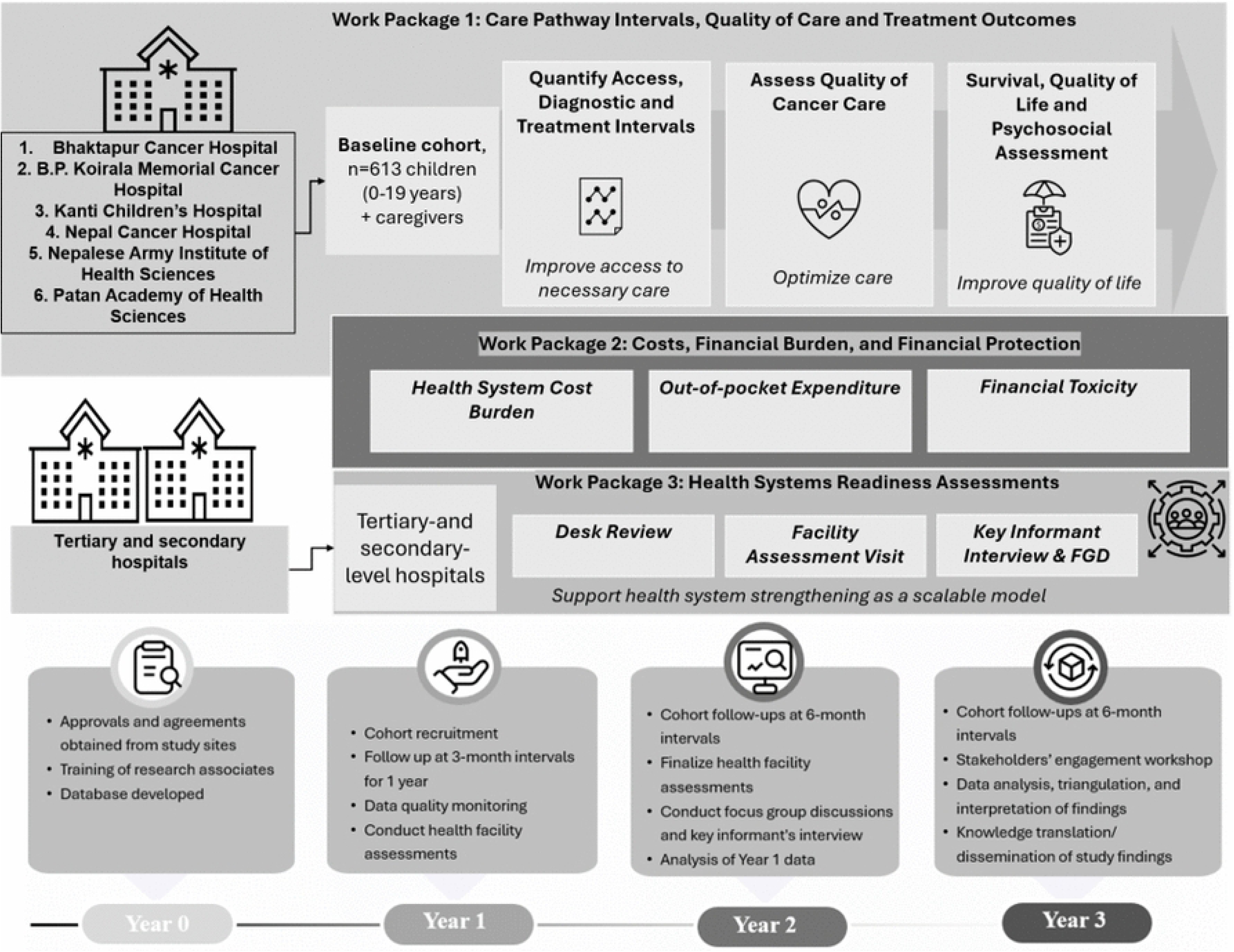
Study settings, work plan and work packages

### Participant eligibility and cohort recruitment

We will seek to recruit consecutive cases of childhood cancer patients and their caregivers registered at six identified hospitals in Nepal that provide care for both urban and rural populations. Cohort recruitment began in January 1, 2026 and will continue until December 2026. To be eligible for inclusion, patients must meet the following criteria: (1) must be children aged 0 to 19 years old with confirmatory cancer diagnosis, (2) present within 12 months of initial cancer diagnosis, (3) able to provide assent by the patients (except for infants and toddlers) and written informed consent by caregiver or legal guardians. Patients who receive cancer diagnosis outside the study facilities or receive partial treatment outside the facility will be eligible for inclusion. Patients who do not meet these criteria or who are present for care due to a recurrence of disease initially diagnosed beyond 12 months from enrolment will be excluded. Participants who provide informed consent and complete a baseline interviewer-administered questionnaire (detailed below) will be followed every 3 months for the first year after diagnosis, and then every 6 months till 3 years after diagnosis.

### Data collection tools and analyses

#### Work package 1: Care Pathway Intervals, Quality of Care and Treatment Outcomes

Baseline interviews of cancer patient/caregivers will be conducted by trained local research assistants using a semi-structured questionnaire. The questionnaire includes patient/caregiver’s sociodemographic characteristics and cancer diagnosis process, including initial symptom recognition, healthcare visits, and details of diagnostic investigations and treatment. Dates of investigations and treatments will be verified with the patient’s medical records. Additional information will be collected on caregivers’ understanding and experiences of their child’s cancer, perceived symptom burden, diagnostic pathways, psychosocial burden, financial toxicity, and quality of life. The questionnaire has been piloted with a representative sample of childhood cancer patients’ caregivers in Nepal. All data collection variables are outlined in Table 1.

**Table 1.**
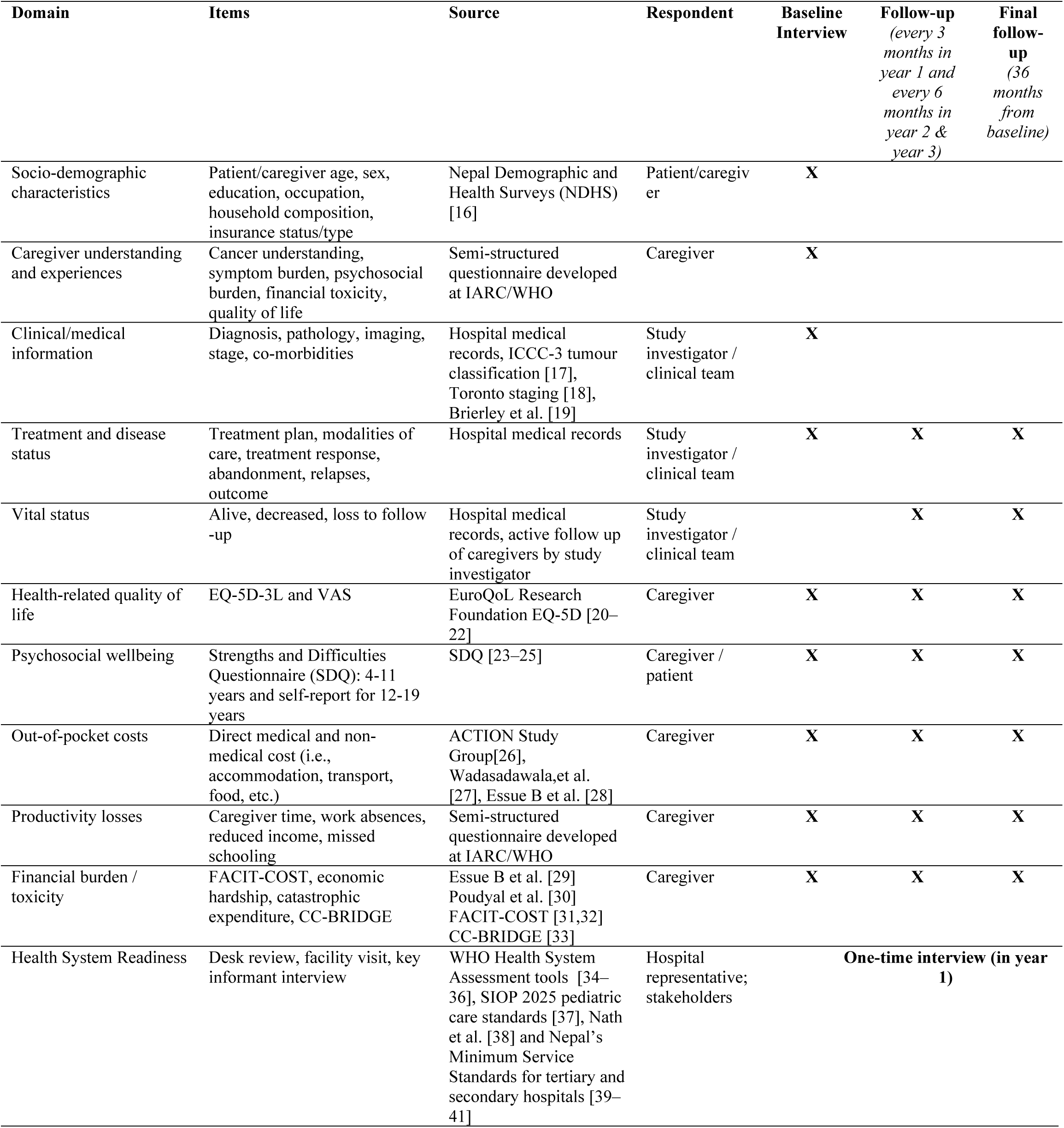
Study tools, variables, and data collection schedule.

Clinical data at the patient level, encompassing pathological and diagnostic information, imaging findings, and documented symptoms, will be systematically abstracted from medical records by locally based research assistants using a standardised case report form. Missing information will be obtained from relevant hospital departments. If a histopathology report is unavailable to confirm a cancer diagnosis for certain solid tumours, diagnosis based on cytology or clinical evaluation supported by radiological or endoscopic findings will be accepted, especially for advanced cancers. For haematological malignancies such as leukaemia, diagnosis will rely on peripheral blood or bone marrow examination findings supported by immunophenotyping results. The International Classification of Childhood Cancer, third edition (ICCC-3) [17] will be used to code tumour diagnosis, and tumor stage will be coded according to Toronto Childhood Cancer Stage Guidelines.[18]

For each patient in the cohort, quality of care will be benchmarked against management regimens from the National Comprehensive Cancer Network, the International Society of Paediatric Oncology Global Health Network Adaptive Management Guidelines, and the resource stratified guidance provided in the ARIA Guide (Adapted Resource and Implementation Application Guide) [42]. Vital status will be confirmed through hospital records or follow-up with clinicians or caregivers. Loss to follow-up will be minimized through collection of multiple contact details and verification through municipal records.

Health-related quality of life (HRQoL) will be assessed using the Nepali version of the proxy administered EQ-5D-5L [20–22], which includes five dimensions: mobility, self-care, usual activities, pain or discomfort, and anxiety and/or depression. Psychosocial strengths and difficulties will be evaluated using the Strengths and Difficulties Questionnaire (SDQ) [23–25] administered as caregiver report for children aged 4–11 years and self-report for those older than 11 years. The SDQ is validated in Nepal and officially been translated into Nepali language [43].

The cohort will be described overall and by cancer type using frequencies and proportions for categorical variables and medians with interquartile ranges for continuous variables, given anticipated non-normal distributions. Intervals from symptom recognition through diagnosis to treatment initiation will be calculated and reported as medians with interquartile ranges, overall and stratified by cancer type, hospital and key sociodemographic characteristics. Associations between patient and caregiver characteristics and interval length will be examined using Chi-squared and Kruskal-Wallis tests for categorical variables and appropriate non-parametric post-hoc pairwise comparisons using the Dunn test with Bonferroni correction. Multivariable associations between explanatory variables and access, diagnostic, and treatment intervals will be assessed using quantile regression models estimating the conditional median (50^th^ percentile) of each interval. Bootstrap methods will be used to obtain standard errors and 95% confidence intervals. Models will adjust for prespecified covariates including cancer type, hospital, child age, sex, and caregiver sociodemographic characteristics. Robust standard errors clustered at the hospital level will be used to account for within-hospital correlation. Results will additionally be reported as adjusted differences in interval length with 95% confidence intervals. Generalized linear models will be used to assess the association of patient’s characteristic and associated determinants of care with HRQoL measures of utility, Visual Analog Scale (VAS) score, and the five EQ-5D-5L domains. Survival at one, two, and three years following confirmatory diagnosis will be estimated using Kaplan-Meier methods. Multivariable Cox proportional hazards models will identify independent determinants of survival. Random effects models will account for hospital clustering, with unadjusted and adjusted hazard ratios reported by cancer type and for all cancers combined. Missing data will be handled using multiple imputation with sensitivity analyses conducted under alternative assumptions.

#### Work package 2: Costs, Financial Burden, and Financial Protection

Financial toxicity will be assessed using the Nepali language version of the FACIT-COST Functional Assessment of Chronic Illness Therapy–Comprehensive Score for Financial Toxicity (FACIT-COST). [31,32] Family costs will be measured using adapted, validated self-reported surveys to capture out-of-pocket costs, changes in household income, financing strategies, caregiving time, and impacts on living standards. We will estimate total out-of-pocket costs, caregiving time costs, catastrophic health expenditures (defined as out-of-pocket spending exceeding 30% of annual income), and cost-related treatment abandonment.

Health system costs (current and projected) will be evaluated using an adapted Childhood Cancers Budgeting Rapidly to Incorporate Disadvantaged Groups for Equity (CC-BRIDGE) tool [33]. We will collect data across 11 costing categories (including personnel, inpatient and outpatient services, pharmacy, pathology, radiation, imaging, surgery, blood services, and administration) that will include real-world data, modeled data, and data from literature. Where necessary, unit costs will be sourced from WHO-CHOICE and adjusted for inflation using the World Bank’s country-specific annual inflation rate [44–46]. Chemotherapy cost estimates will incorporate treatment intensity, survival rates, and patient weight to support forecasting and resource allocation.

We will analyse annual family out-of-pocket costs using descriptive statistics overall and by site, with Chi-squared tests and t-tests used to examine differences across groups. Disaggregated costs and their drivers will be examined using a two-part regression model, a logistic component to estimate the probability of incurring any expenditure and an ordinary least squares component to model the level of expenditure among those with positive costs, to account for the zero-inflated distribution typical of health care cost data. Health system costs will be estimated from one year of cost data and projected over five years using Nepal’s annual inflation rate and anticipated patient volume growth, aligned with actual and projected GDP. Sensitivity analyses will generate upper and lower bounds to projected costs using anticipated changes in survival rates. Cost projections will be modelled under defined scenarios of health insurance coverage to estimate corresponding rates of catastrophic health expenditure and medical impoverishment for each scenario.

#### Work package 3: Health Systems Readiness Assessment

Data for the health system readiness assessment will be gathered from three primary sources: (1) a comprehensive desk-based review, (2) in-person facility assessments conducted by study investigators, and (3) key informant interviews with multi-level stakeholders. The tool for desk-review will be adapted from the WHO Health System Building Blocks [34], which captures information on national cancer control strategies, including legislation, protocols, program evaluations, quality assurance mechanisms, and other reports from the Ministry of Health. This review will be supplemented by available peer-reviewed and grey literature. Following the desk-review, on-site facility assessments in selected secondary and tertiary hospitals and stakeholder consultations will be conducted. The tool for health facility assessment will be adapted from the WHO Harmonized Health Facility Assessment (HHFA) [35], Standards for Paediatric Oncology Units in low- and middle-income countries developed by the International Society of Paediatric Oncology (SIOP) [37], facility-level assessment checklist developed by the Indian Council of Medical Research (ICMR), India [38], WHO list of priority medical devices for cancer management [36], and the minimum services standard (MSS) checklists developed for quality improvement of tertiary and secondary level hospitals by the Ministry of Health, Nepal [39–41]. Following the facility visits, challenges and gaps identified will be explored through multiple focus group discussions separately with pediatric oncologists, nurses, and patients’ caregivers. In addition, key informant interviews (KII) will be conducted with focal persons from relevant divisions and departments of the Ministry of Health on childhood cancer policy, strategies, and planning including laboratory and diagnostics services, with Department of Drug Administration (DDA) and Health Management Information System (HIMS) for procurement and supply of childhood cancer medicines and equipment and data recordings, with finance department for budget allocation and subsidies, and with patient representative group/organization for patient needs and/or expectations. The interviews are expected to last approximately 90-120 minutes and will follow a co-developed semi-structured guide based on the WHO health system building blocks [34], which will capture barriers and facilitators to high-quality, timely diagnosis and treatment, and survivorship care for childhood cancer patients. Interviews will be recorded and transcribed.

Findings from the desk review, facility assessments, in-depths interviews and key informant interviews will be triangulated and synthesized into a SWOT analysis that will highlight the strengths, weaknesses, opportunities, and threats related to delivery of childhood cancer care in Nepal. The synthesis of various techniques will reflect both the documentary evidence and real-world implementation realities. Data will be analysed descriptively using tracer items adapted from the health facility assessment. Item-level findings will be summarised as frequencies and proportions and grouped into prespecified domains relevant to childhood cancer care, such as governance, workforce, diagnostics, medicines, equipment, referral systems, and information systems. Domain-specific readiness scores will be calculated as the proportion of required tracer items available, expressed as percentages, and compared across facility types and levels of care.

Qualitative data from facility discussions and key informant interviews will be analysed thematically and triangulated with quantitative findings to provide an integrated assessment of health system readiness to produce a more policy-relevant interpretation for improving childhood cancer care in Nepal. However, the assessment will not include evaluation of the clinical management of childhood cancer provided at the time of facility visit.

#### Sample size considerations

We will enroll approximately 619 newly diagnosed childhood cancer patients. The sample size was calculated based on the ability to detect a clinically meaningful absolute difference of 5.5% in survival compared with the historical childhood cancer survival estimate in Nepal. Assuming a baseline survival rate of 30% (p0 = 0.30), an expected survival rate of 35.5% (p1 = 0.355), a two-sided significance level of 5% (Zα/2 = 1.96), and 80% power (Zβ = 0.84 for 80% power, corresponding to z-value at 80th percentile in standard normal distribution), the required sample size was estimated to be 557 patients. To account for the potential of 10% lost to follow-up, the inflated sample size required is n_2_=557/ (1-0.10) = 619. Assuming a 30% survival rate, approximately 434 deaths are expected during follow-up. This number of outcome events will provide sufficient statistical power to estimate survival outcomes using multivariable regression models and identify clinically meaningful differences in survival associated with demographic, clinical, treatment-related, and health system characteristics.

#### Data management

Patient and caregiver baseline data will be collected first on paper-based questionnaires. Participant study data will be entered and stored in IARC/WHO’s Research Electronic Data Capture (REDCap) database, with no personal identifiable health information. A REDCap-generated unique study identifier will be assigned to each participant. All participant-identifying information, including name, hospital number, and contact information, will be stored only in facility records and will not be included in stored study data. De-identified data will be securely shared with all study team members via REDCap to ensure equal access to the data and support subsequent analysis. All complete questionnaires and data collection forms will undergo periodic checks by the investigator and the study team to ensure accuracy and completeness.

## DISCUSSION

This study will provide foundational evidence to inform Nepal’s cancer control planning, strengthening childhood cancer care and cancer registration, and inform comparable efforts in other low- and middle-income countries (LMICs). Enhancing hospital-based and national data systems and aligning them with global datasets supported by the International Agency for Research on Cancer, has implications beyond surveillance as robust data systems are essential for resource allocation, workforce planning, and equity monitoring [1,47]. Implications of our findings will align well to benchmark the progress on WHO’s GICC initiative and several targets of Sustainable Development Goals [1].

This study will address a critical gap in the literature on financial protection for childhood cancers [48], including evidence on household economic burden and its implications for treatment abandonment and survival. Our findings will emphasize equity, financial protection, and health system strengthening in childhood cancer care in Nepal. By examining cost structures, financing mechanisms, and system-level constraints, this work advances understanding of how financial risk protection can be operationalized within national cancer control strategies.

### Ethical considerations

Ethics approval for this study was obtained from the Nepal Health Research Council’s national ethics committee. Approval was also received from the Institutional Review Board of the International Agency for Research on Cancer, and the Research Ethics Board at the University of Toronto, and the relevant institutional review committees at all six participating hospitals in Nepal. All procedures comply with international ethical standards for research involving human participants and with national regulatory requirements.

Written informed consent will be obtained from caregiver or legal guardians of all participating children, with assent obtained from children. Participant confidentiality will be strictly maintained through de-identification of data, secure digital storage, and restricted access to study databases. Data sharing agreements between institutions will govern the transfer, storage, and use of data in alignment with applicable data protection policies.

An independent study advisory committee composed of national and international experts, clinicians, policymakers, and lived experience experts will provide ethical and scientific oversight throughout the study period. This committee will monitor study conduct, advise on emerging ethical issues, and support responsible dissemination of findings to ensure that results are translated into policy and practice in an equitable and context-appropriate manner.

### Dissemination plan

We will disseminate findings among various target audiences including general population, patient representatives, clinical societies, local government and non-governmental organizations, and the Ministry of Health. These findings will support the development of a national database for childhood cancer and identify priority areas to improve outcomes in childhood cancer, identify opportunities to reduce family’s financial hardship, and to identify areas to strengthen cancer care delivery capacity of health system in Nepal. The findings from this study will be shared through open-access publications, conference presentations, lay summaries, and a multi-stakeholder dissemination meeting. A final study report as a ‘strategy document’ will be submitted to the Ministry of Health and Population (MoHP), Nepal.

## Acknowledgement

The authors would like to thank the respective departments and focal persons of all involved hospitals that are helping in the smooth extraction of the medical records of the patients. We would also like to thank all patients and caregivers who are voluntarily participating in this study. The authors would like to thank Caitlin Laska for her support in drafting the protocol.

## AUTHORS’ CONTRIBUTIONS

D.S., A.M., B.E., P.B. — Conceptualization, Funding acquisition, Methodology, Writing – review & editing. All authors reviewed and edited the manuscript draft for finalization.

### Disclaimer

Where authors are identified as personnel of the International Agency for Research on Cancer/World Health Organization, the authors alone are responsible for the views expressed in this article and they do not necessarily represent the decisions, policy, or views of the International Agency for Research on Cancer/World Health Organization. Martiniuk was funded (to Dec 31, 2025) by an Australian National Health and Medical Research Council Investigator Grant (NHMRC #1195086).

### Data Availability

All relevant data from this study will be made available upon study completion.

### Funding

The study is funded by the Canadian Institutes of Health Research (CIHR - 542698). The funder of the study had no role in study design, data collection, data analysis, data interpretation, or writing of the report. DS, BE, AM, MI, PB will have full access to all data. The corresponding author has the final responsibility to submit for publication.

